# Clinical Characteristics and Severity of COVID-19 Disease in Patients from Boston Area Hospitals

**DOI:** 10.1101/2020.07.27.20163071

**Authors:** Hesam Dashti, David W. Bates, Elise Roche, Julie Fiskio, Samia Mora, Olga V. Demler

**Affiliations:** Division of Preventive Medicine, Brigham and Women’s Hospital and Harvard Medical School, Boston, MA, USA; Division of Preventive Medicine, Brigham and Women’s Hospital, USA; Division of Preventive Medicine, Brigham and Women’s Hospital, Boston, MA, USA; Information Systems, Partners HealthCare System, Wellesley, MA; Division of Cardiovascular Medicine, Brigham and Women’s Hospital and Harvard Medical School, Boston, MA, USA

**Keywords:** COVID-19, SARS-CoV-2, medication use, comorbid conditions

## Abstract

We summarize key demographic, clinical, and medical characteristics of patients with respect to the severity of COVID-19 disease using Electronic Health Records Data of 4,140 SARS-CoV-2 positive subjects from several large Boston Area Hospitals. We found that prior use of antihypertensive medications as well as lipid lowering and other cardiovascular drugs (such as direct oral anticoagulants and antiplatelets) all track with increased severity of COVID-19 and should be further investigated with appropriate adjustment for confounders such as age and frailty. The three most common prior comorbidities are hyperlipidemia, hypertension, and prior pneumonia, all associated with increased severity.

## Background

The incidence of COVID-19 in Massachusetts has been near exponential, ranking third nationwide with more than 66,000 confirmed case. Partners HealthCare, which includes several large Boston-area hospitals, has reported 4,140 COVID-19 patients as of April 18, 2020. We summarize key demographic, clinical, and medical characteristics of these patients with respect to the severity of COVID-19 disease.

## Methods

The Partners Electronic Health Record (EHR) contains outpatient and inpatient records for >4 million patients mostly from greater Boston, Massachusetts. The patient population in this study includes all patients (N=4,140) with at least one positive or presumptive positive COVID-19 test result^1^ documented in the Partners EHR between December 1, 2019 and April 18, 2020. The study was approved by the Partners Institutional Review Board. Laboratory, clinical, and medical records (April 1, 2015-April 18, 2020) were obtained from the Partners Research Patients Data Registry (RPDR) and the Enterprise Data Warehouse (EDW)^2-4^. All data was updated daily, except for prior conditions which are updated annually using a machine learning algorithm^4^. Income data was linked to the 2018 US Census. Medication use was coded as ever in 2015-2020. Severity was defined in five mutually exclusive categories (I) outpatients only, (II) evaluation in an emergency department without admission, (III) inpatient admission not requiring intensive care unit (ICU) care, (IV) inpatient admission requiring ICU care, and (V) deceased.

Two-sided tests of trend were performed unless otherwise noted. Python and R software languages were used. All data were obtained from Electronic Health Records Repository Maintained by Brigham Mass General HealthCare Systems in full compliance with the IRB protocols and met all data access requirements. Obtaining consent from all individuals would make the study challenging logistically, financially and scientifically, especially due to the urgent nature of the pandemic.

## Results

Out of 4,140 COVID-19 patients (55% women, median age 52 years (interquartile range: 36-65)), two thirds (N: 2,759) were outpatients, including 795 (19% of the COVID-19 patients) who were evaluated in an outpatient emergency department. Of the 1,194 inpatients, 619 (15% of the COVID-19 patients) were admitted to a Partners hospital but not to the ICU, while 575 (13.9%) were admitted to the ICU, and 187 (4.5%) died in-hospital. Older age, male sex, and comorbidities were more prevalent according to COVID-19 severity (Table). Prevalence of current/ever smoking was greater by severity (p-value < 2.2e- 16), and this group accounted for 58.3% of the deceased patients. Median household income per zip code had nonlinear association with severity. Table shows comorbidities and medication use according to severity. The most prevalent comorbidities among deceased patients were hyperlipidemia, hypertension, prior pneumonia (p-values < 2.2e-16 from the test of trend). Multiple other comorbidities also tracked with the disease severity. Use of angiotensin converting enzyme inhibitors or angiotensin receptor blockers was also greater by severity, but so was use of cardiovascular and other medications including other antihypertensives, statins, antiplatelets, direct oral anticoagulants, antidiabetic medications and vitamin D3.

**Table.**
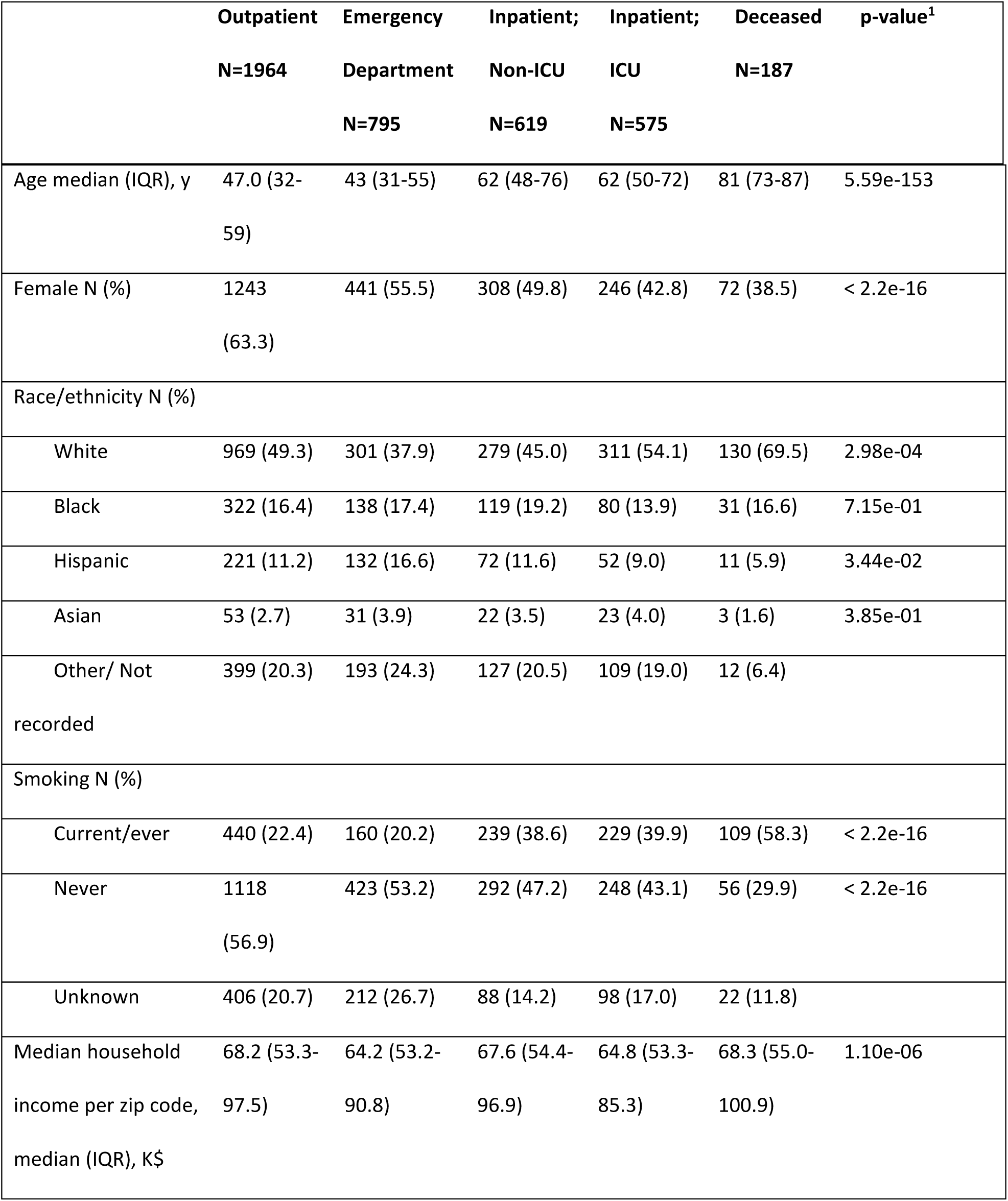
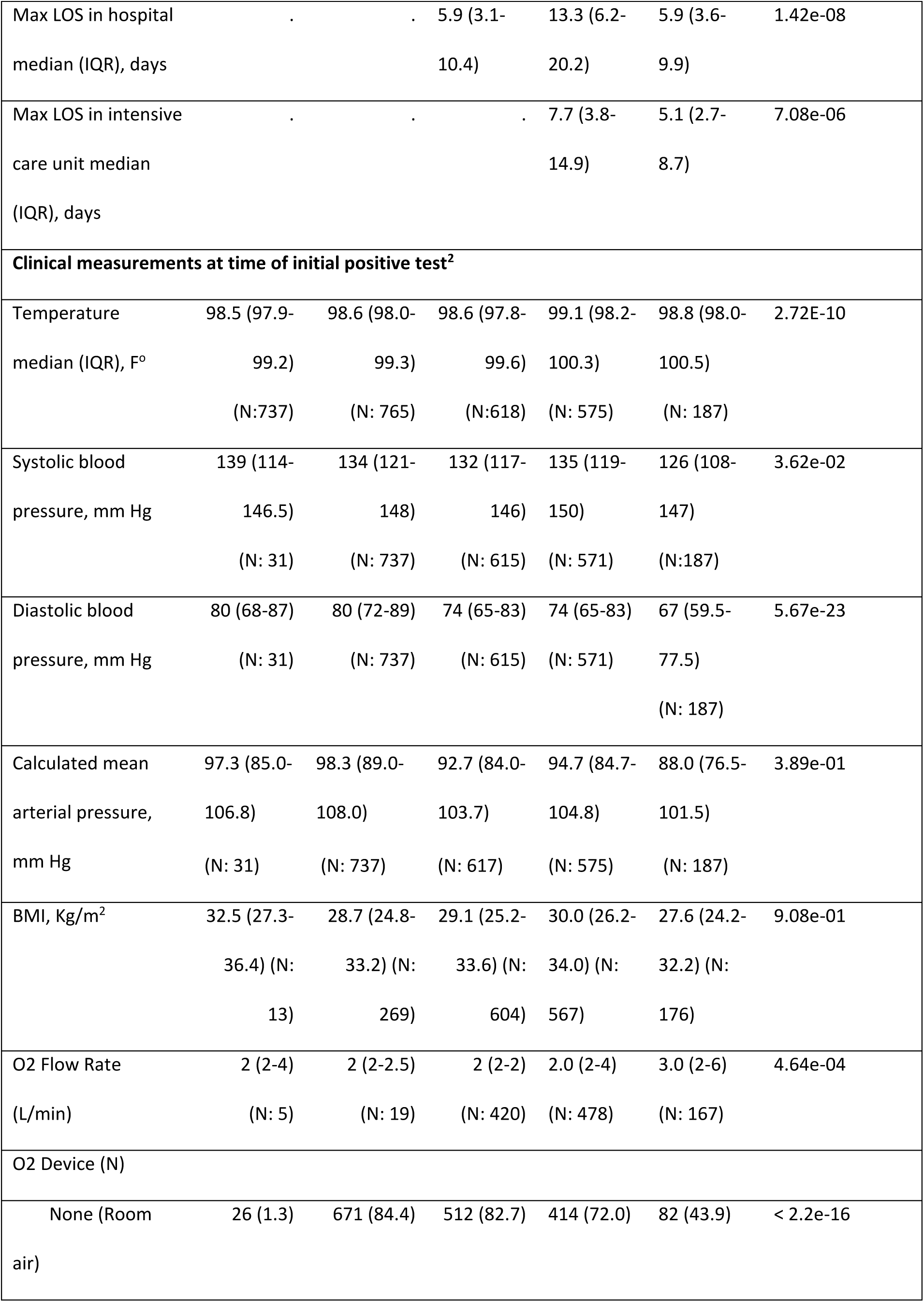
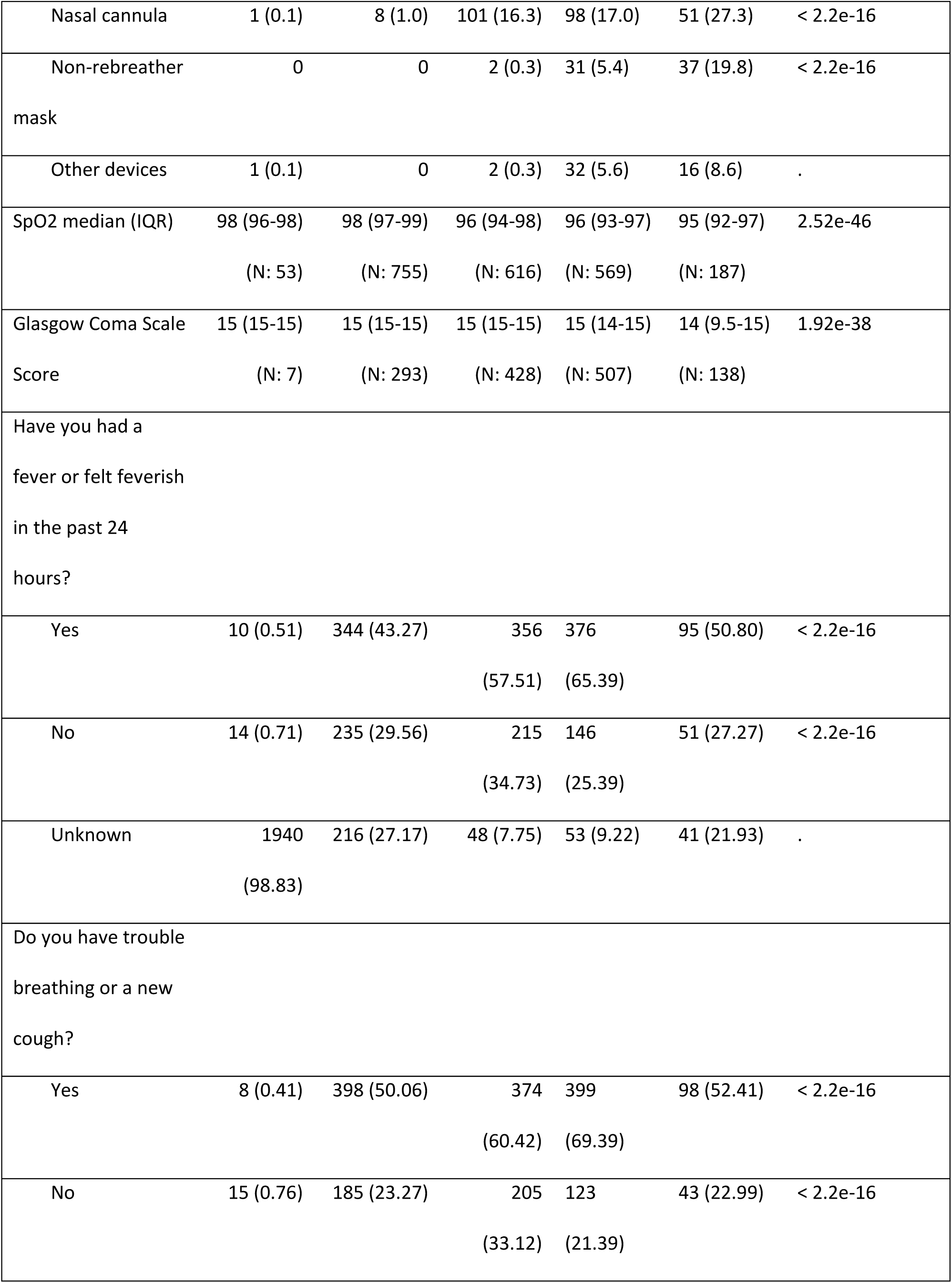
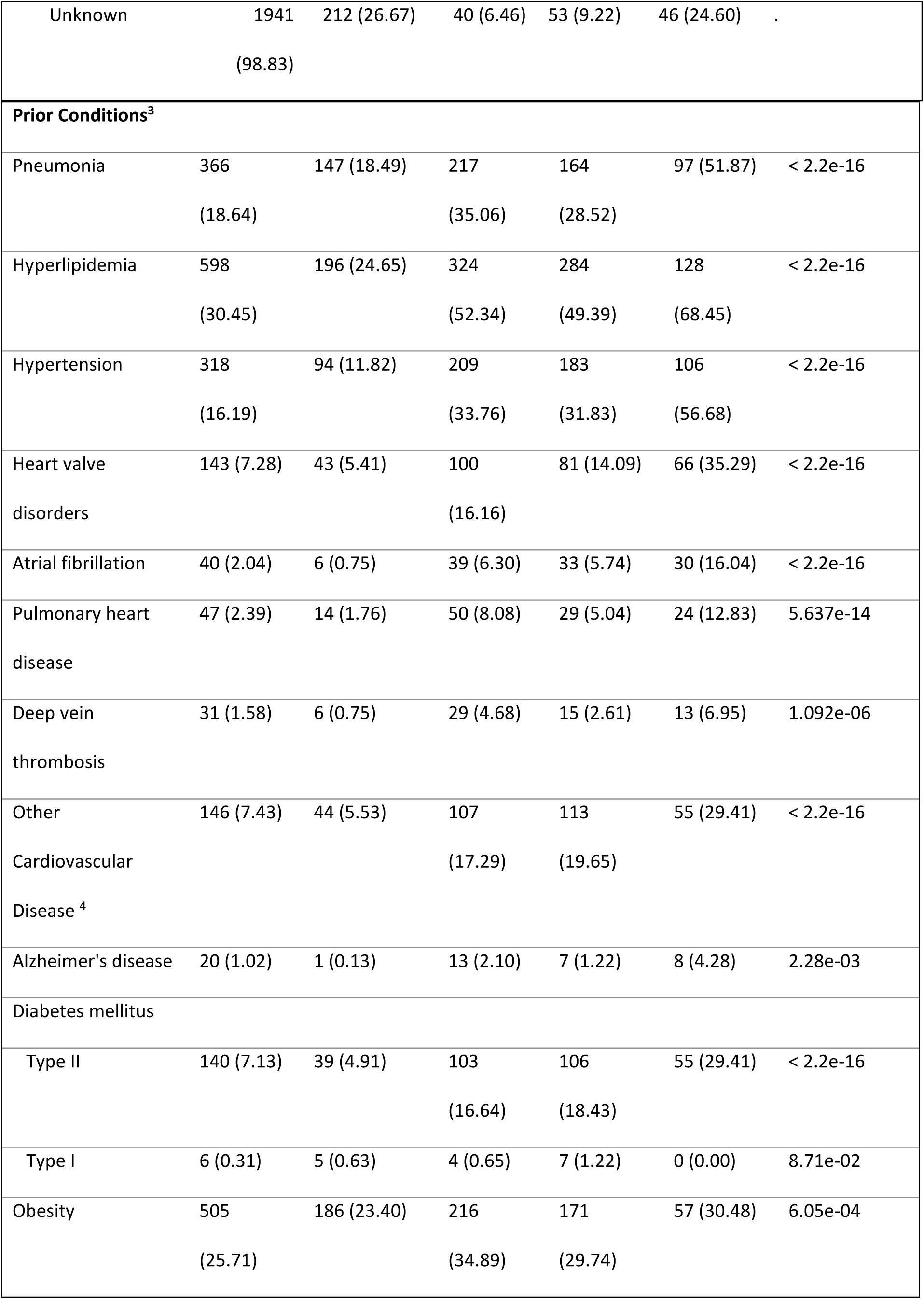
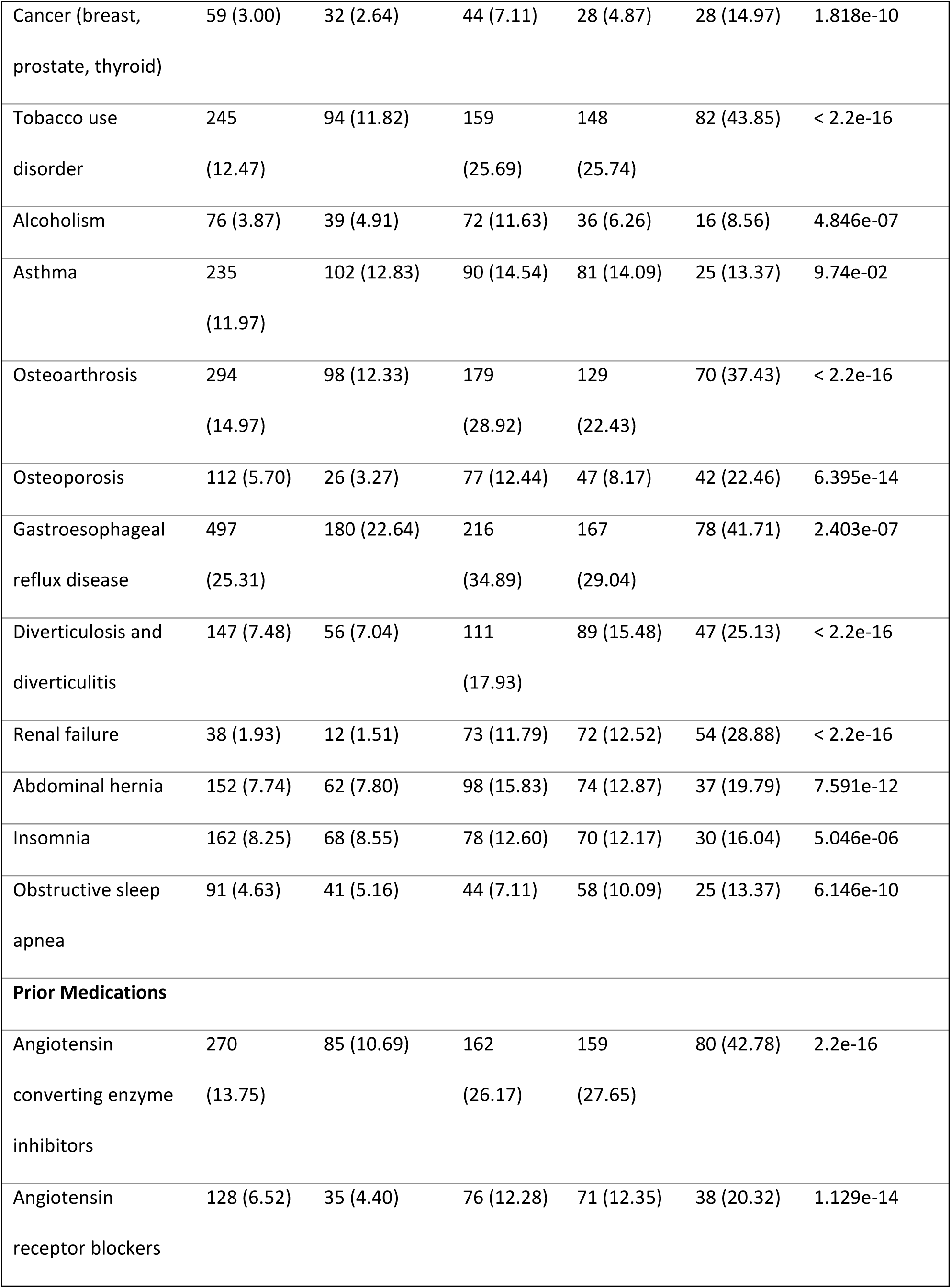
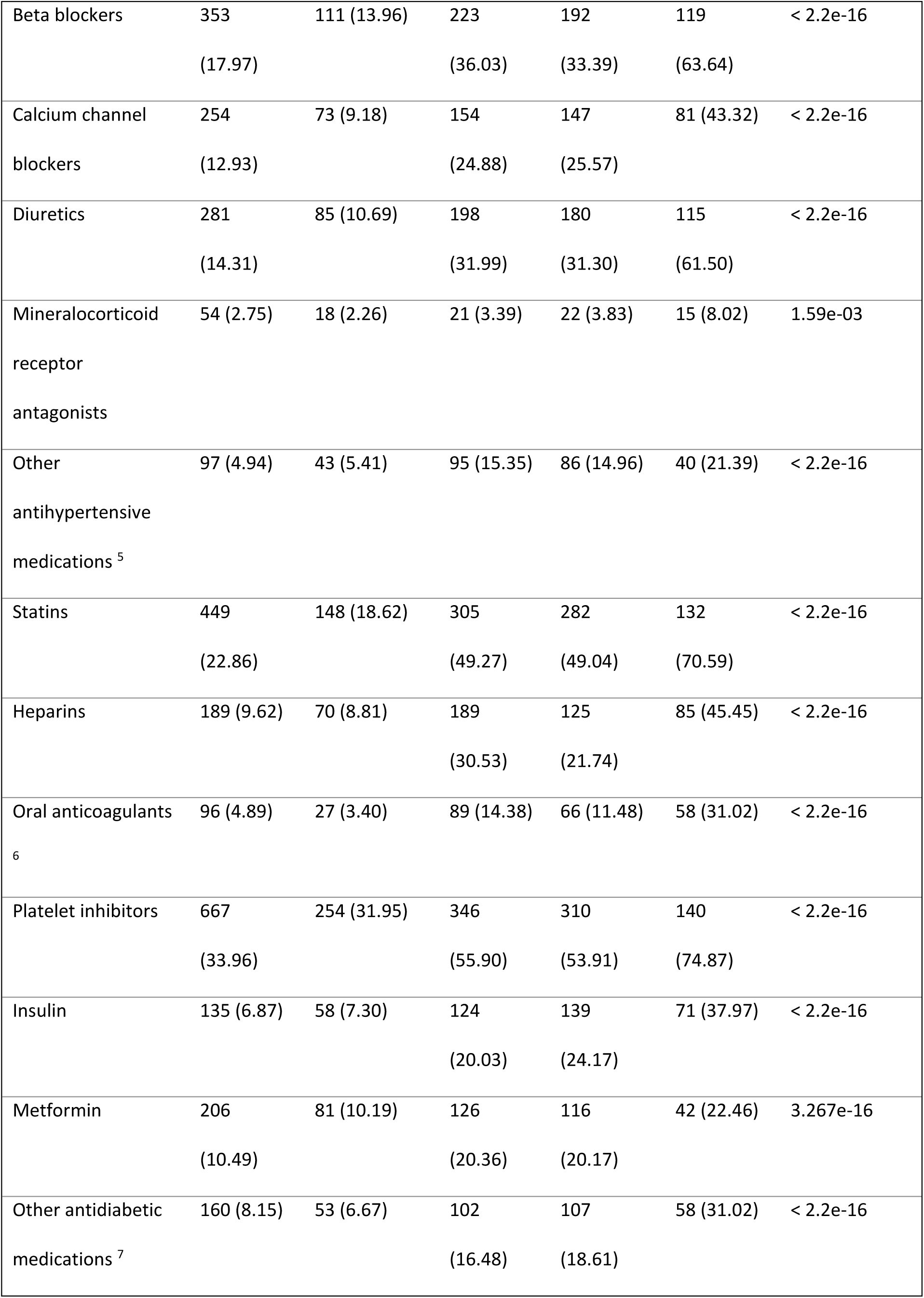
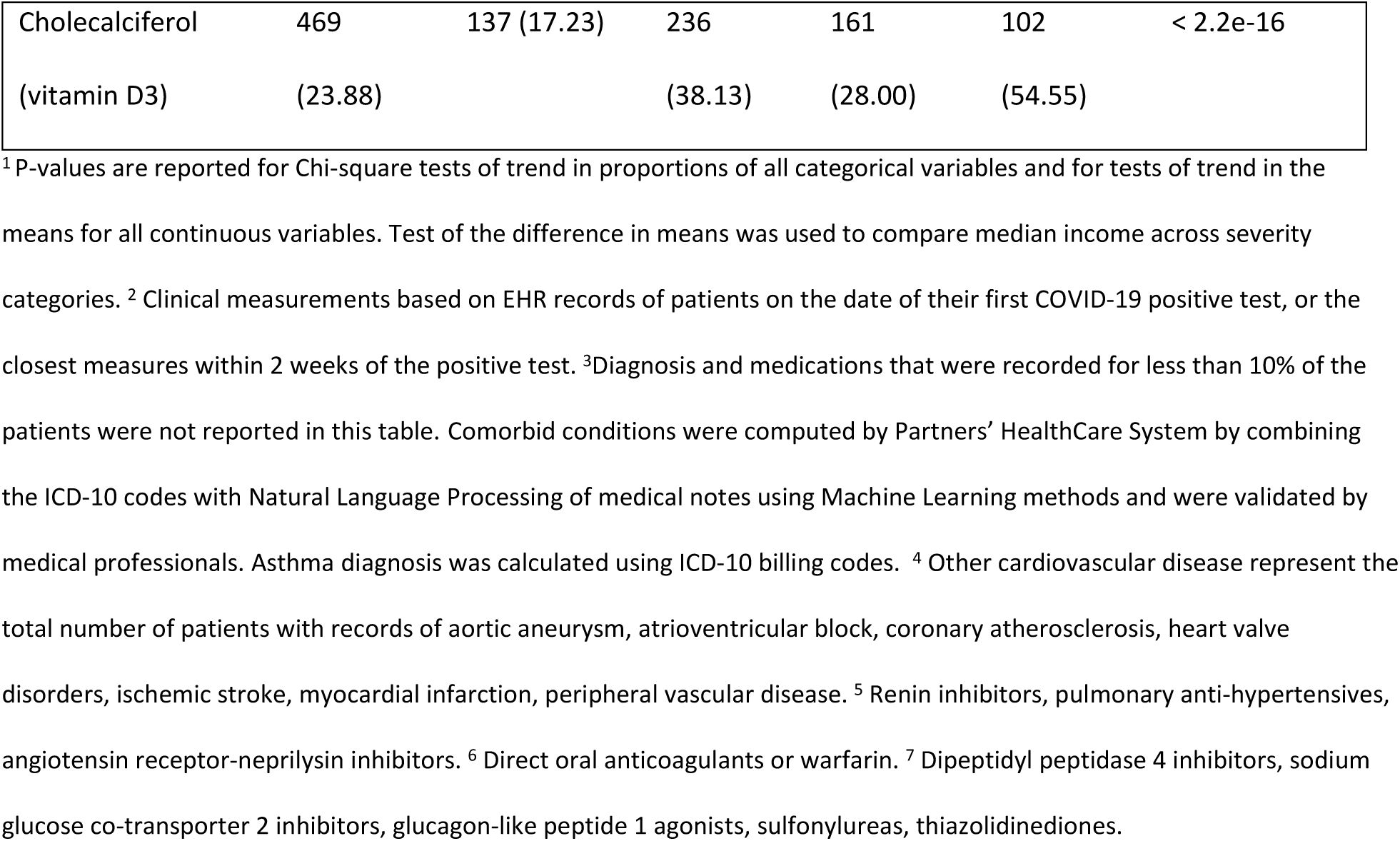
Demographic characteristics, clinical measurements, prior conditions and medication use by severity of COVID-191.

## Discussion

In this large greater Boston study of 4,140 patients, multiple comorbidities and greater use of medications tracked with the severity of COVID-19. The data sources used in this study were limited to the use of the EHR. We found that prior use of antihypertensive medications as well as lipid lowering and other cardiovascular drugs such as direct oral anticoagulants and antiplatelets all tracked with increased severity of COVID-19 and should be further investigated with appropriate adjustment for confounders^5^. Common prior comorbidities including hyperlipidemia, hypertension, and pneumonia also tracked with increased severity.

## Data Availability

Partners EHR repositories are controlled-access databases and cannot be posted publicly due to a superseding Partners Healthcare policy that restricts access to internal researchers to protect privacy of patients.

## Footnotes

Authors of this manuscript have no conflicts of interest to declare.

